# Clinical, Prognostic and Biological Features of High-Risk Cardiometabolic Phenotype: the REMODEL Study

**DOI:** 10.64898/2026.01.16.26344303

**Authors:** Adam AR Muhammad, Jia Kai Chuan, Aisyah Latib, Jennifer A Bryant, Vivian Lee, Redha Boubertakh, Thu-Thao Le, Calvin WL Chin

## Abstract

**Background:** Conventional cardiometabolic risk markers (blood pressure, glucose, lipids) incompletely capture biological vulnerability in hypertension. We aimed to identify distinct risk phenotypes and define their clinical, prognostic, and proteomic features.

**Methods:** REMODEL is a prospective observational cohort of asymptomatic adults with essential hypertension who underwent 24-hour ambulatory blood pressure monitoring and standardized cardiovascular magnetic resonance (CMR). Among 83 candidate predictors, outcome-informed feature selection identified four variables with best discrimination: NT-proBNP, Romhilt–Estes ECG score, and CMR-derived indexed interstitial and myocyte volumes. Unsupervised clustering using the KAMILA algorithm (without outcome labels) determined two clusters. The primary outcome was a composite of acute coronary syndromes, first heart failure hospitalization, stroke, and all-cause mortality. Proteomics quantified 192 cardiovascular-related proteins (Olink CVD II/III).

**Results:** Of 885 participants, two clusters emerged: low-risk (n=787) and high-risk (n=98). Over 60 [37,73] months, the high-risk cluster had markedly worse event-free survival (log-rank P<0.001) and remained independently associated with outcomes (adjusted HR 8.89, 95% CI 4.75–16.65). High-risk individuals were younger, more often male, had greater visceral adiposity, worse renal function, higher 24-hour blood pressures, and adverse remodeling on CMR/biomarkers. Proteomics identified 26 enriched proteins implicating myocardial stress, extracellular matrix remodeling/fibrosis, immune activation, apoptosis, and vascular dysfunction.

**Conclusions:** Multimodal clustering reveals a high-risk hypertensive phenotype with distinct structure–biology coherence and substantially elevated long-term risk, supporting integrated imaging–biomarker–proteomic approaches for refined stratification.

---

Traditional risk stratification in cardiometabolic diseases relies on markers like blood pressure (BP), glucose, and lipid levels. While informative, these parameters fail to capture the full biology underpinning cardiometabolic risk. Notably, nearly 30% of hypertension-related health burden occur in patients with apparently well-controlled BP^1^, highlighting the limitations of conventional metrics and the need to uncover mechanistic pathways driving vulnerability. In this study, our objectives are to identify distinct risk profiles using the KAMILA algorithm^2^, and characterise the clinical, prognostic and biological features that define high-risk hypertensive phenotype.

The **REMODEL** (Response of the myocardium to hypertrophic conditions in the adult population; clinicaltrials.gov unique identifier: NCT02670031) is a prospective, observational cohort of asymptomatic individuals with essential hypertension^3^. Exclusion criteria included secondary causes of hypertension, known cardiovascular disease, inherited cardiomyopathies, atrial fibrillation, or contraindications to gadolinium or CMR. All participants underwent 24 ambulatory BP monitoring (OnTrak 90227, SpaceLabs Healthcare) and standardized cardiovascular magnetic resonance (CMR; imaging details provided as online supplemental data), with de-identified images centrally analyzed at the **National Heart Research Institute of Singapore CMR Core Laboratory** using standardized protocols^4^.

Measurement of 192 serum proteins related to cardiovascular diseases were performed using 2 commercially available multiplex immunoassays (Olink Target Cardiovascular Disease II and III, Olink Proteomics, Uppsala, Sweden). This proteomic analysis enabled us to investigate molecular pathways that may contribute to adverse outcomes observed in high-risk hypertensive phenotype. The primary outcome was a composite of first occurrence hypertension-related adverse events: acute coronary syndromes, first heart failure hospitalization, strokes and all-cause mortality. Patients were followed until December 2024, with data censored at the last known event-free date for individuals lost to follow-up. Clinical events were adjudicated through medical record review using predefined criteria previously published^3^. The cause of death was ascertained from the National Death Registry.

Continuous variables were summarized as mean ± SD or median (IQR) where appropriate, and categorical variables as counts and percentages. Between-group comparisons were performed using t-tests or Mann-Whitney U tests for continuous data, and χ^2^ tests for categorical data. Event-free survival was assessed with Kaplan-Meier analysis with log-rank testing. All analyses were performed in R (v4.4.1) and Python (v3.12.4), with a two-sided P < 0.05 considered as statistically significant.

Of the 885 individuals with hypertension, 21% had diabetes mellitus and 49% had dyslipidemia. The mean body mass index was 26.4±4.5kg/m^2^ and 24-hour BP was 131±14/79±10 mmHg. Among the 83 candidate predictor variables assessed, four were selected for their superior discriminative performance: N-terminal pro-Brain Natriuretic Peptide (NT-proBNP), Romhilt-Estes electrocardiographic score, and indexed interstitial and myocyte volumes—both derived from cardiovascular magnetic resonance (CMR). This combination yielded an area under the receiver operating characteristic curve (AUC) of 0.76 (95% CI: 0.68–0.83; P<0.001) for predicting adverse outcomes. In contrast, the best-performing clinical predictor, 24-hour systolic BP, ranked eighth with an AUC of 0.66 (95% CI: 0.57–0.74; P<0.001). The silhouette method identified two optimal clusters, with the highest silhouette score of 0.76. Unsupervised clustering of the cohort was performed using the KAMILA algorithm, and the two clusters demonstrated clear separation in principal component analysis (Figure 1). Predictor selection was outcome-informed to enrich for clinically meaningful phenotypes, while cluster assignment itself was performed without outcome labels. This hybrid approach was designed to enhance clinical interpretability but may inflate associations between cluster membership and outcomes; and should therefore be considered hypothesis-generating.

**Figure 1.**
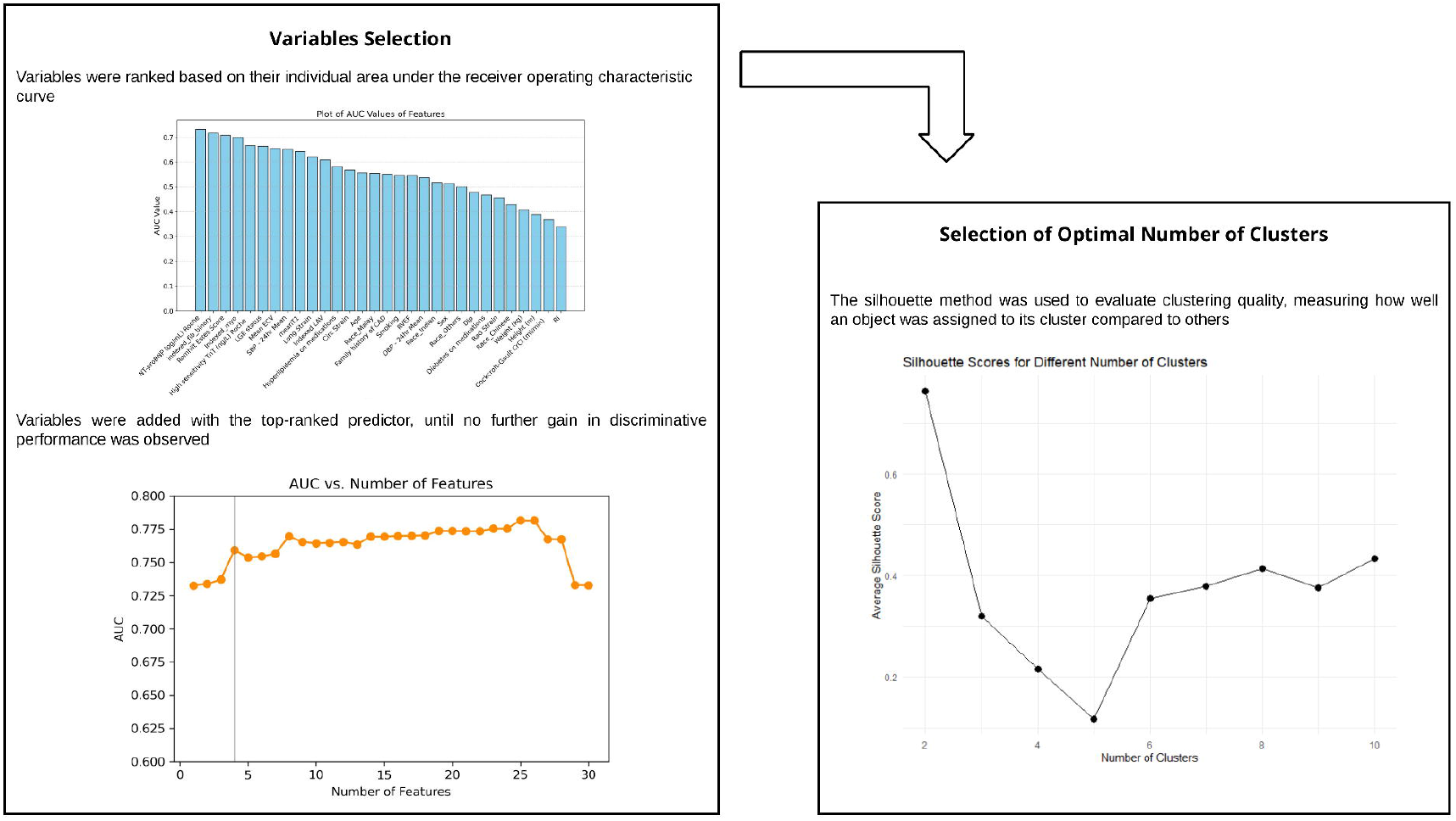
Determining Number of Clusters and Cluster Assignment. Four predictor variables were selected for their superior discriminative performance and the optimal number of clusters was two. KAMILA clustering of the cohort demonstrated clear separation in principal component analysis

These clusters included a low-risk (n=787) and a high-risk (n=98) group. Individuals in the high-risk group were more likely males, younger (54±14 versus 58±10 years, P<0.001), worse renal function and a greater visceral adiposity (200.0 [148.1, 297.3] versus 160.7 [103.1, 232.0] cm^2^, P=0.007) and higher 24-hour blood pressures (144±16/84±13 versus 129±13/79±9 mmHg). Individuals in the high-risk cluster also had more adverse features of cardiac remodeling on both CMR and circulating biomarkers (Figure 2a). Of note, individuals in both clusters had similar subcutaneous adiposity (P=0.291). During a follow-up period of 60 [37, 73] months, the high-risk cluster experienced significantly worse outcomes (Figure 2b; P<0.001); and remained significant even after adjusting for potential confounders (HR 8.89 [95% CI 4.75–16.65]; P < 0.001).

**Figure 2.**
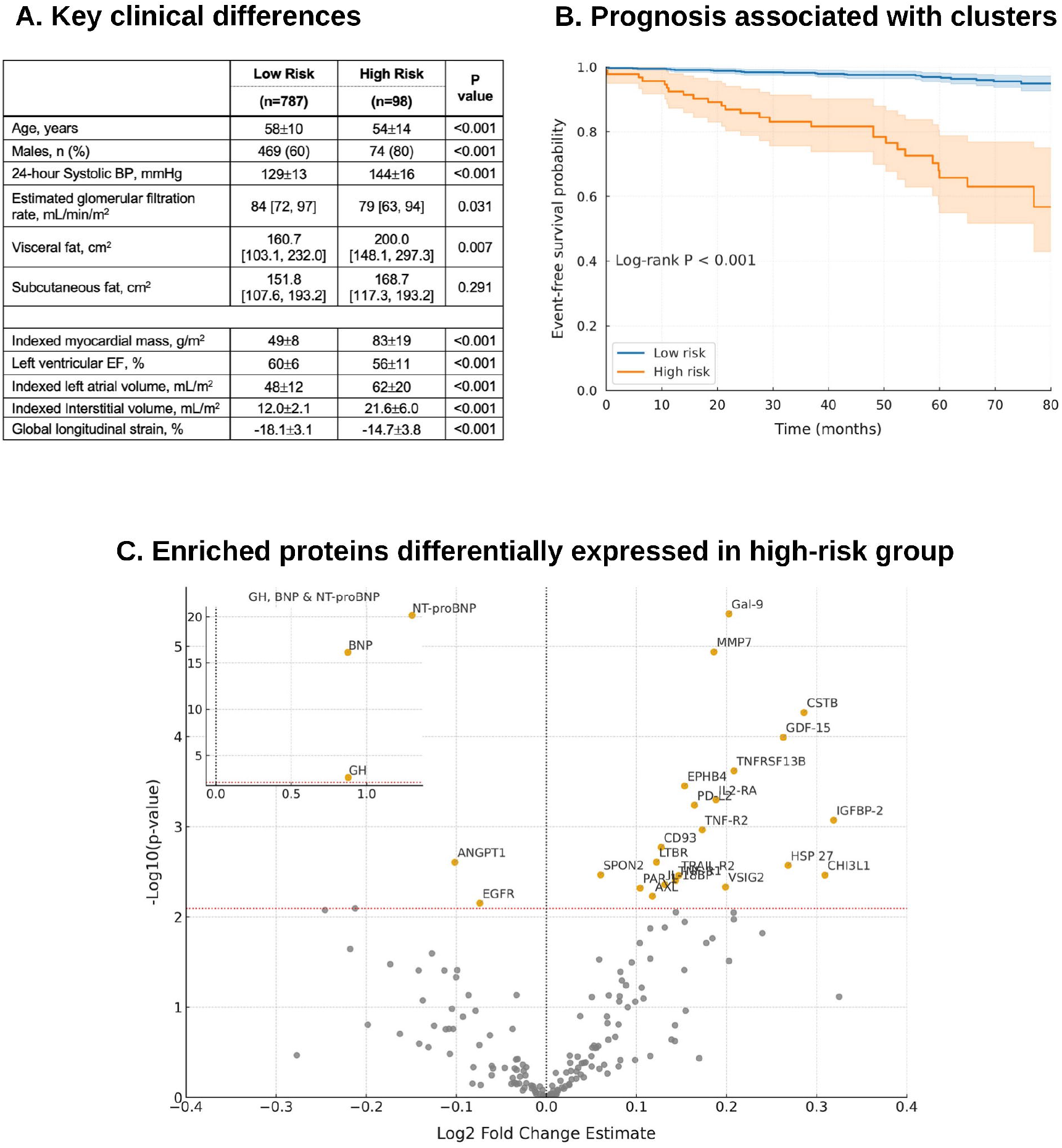
Clinical, prognostic and biological features associated with high-risk cluster. Those in the high-risk cluster were younger, more likely males, greater visceral adiposity, had worse renal function and higher 24-hour blood pressures (Panel A). High-risk cluster was also associated with significantly worse prognosis (Panel B). Proteomic profiling identified 26 circulating proteins significantly enriched in the high-risk cluster (Panel C). Abbreviations: BP blood pressure; EF ejection fraction.

Proteomic profiling revealed 26 circulating proteins significantly enriched in the high-risk hypertensive cohort (Figure 2c). Classical cardiac stress biomarkers (BNP and NT-proBNP) were prominently elevated, consistent with myocardial stretch and neurohormonal activation. Proteins mediating extracellular matrix turnover and fibrosis (MMP7, GDF-15, CSTB and HSP27) were also significantly enriched. A broad set of immune regulators and cytokine mediators (Galectin-9, TNF-R1, TNF-R1, TRAIL-R2, TNFRSF13B, IL2-RA, IL-18BP, PD-L2, CD93, CHI3L1, LTBR, and VSIG2) suggested heightened inflammatory activity, immune checkpoint modulation, and pro-apoptotic signalling. In parallel, vascular signaling and angiogenic proteins (EPHB4, ANGPT1, AXL, EGFR, PAR-1, and SPON2) implicated endothelial dysfunction, angiogenesis, and thrombo-inflammatory processes.

The identification of the four key predictors underscore the importance of integrating multimodal data, including biomarkers and advanced imaging, for risk assessment. NT-proBNP, a well-established marker of cardiac stress, has been consistently linked to adverse cardiovascular outcomes. Beyond this, proteomic profiling provided additional mechanistic insights into the high-risk hypertensive phenotypes. Twenty-six proteins were significantly enriched, spanning pathways of myocardial stretch, extracellular matrix remodelling, inflammation, apoptosis and vascular dysfunction. Taken together, these circulating signatures complement structural insights from CMR-derived measures of interstitial and myocyte volumes, which reflect microstructure and subclinical remodelling; and electrocardiographic findings, such as the Romhilt-Estes score which capture left ventricular hypertrophy^5^. Proteomic enrichment of natriuretic peptides therefore reflects internal biological coherence rather than independent validation of cluster distinctiveness.

Visceral adiposity emerged as a key clinical feature of the high-risk cluster. Despite similar levels of subcutaneous fat, individuals in this group exhibited substantially greater visceral fat burden, a phenotype strongly associated with systemic inflammation, insulin resistance, and adverse cardiometabolic outcomes. Importantly, the high-risk cluster was paradoxically younger, suggesting that adverse cardiovascular risk in hypertension may be driven by early-onset metabolic and myocardial vulnerability rather than chronological aging alone. This finding highlights limitations of conventional risk stratification tools, which often underestimate lifetime risk in younger individuals with adverse biological profiles.

This study is novel in demonstrating that unsupervised risk clustering not only improves prediction but also reveals disease biology, linking clinical features with molecular signatures to refine risk assessment. However, it has several limitations. The modest high-risk sample limits generalizability. Although two clusters were optimal by silhouette score, finer sub-phenotypes may emerge in larger cohorts. Use of all-cause mortality broadened capture of hypertension-related fatal outcomes but reduced mechanistic specificity compared with cardiovascular mortality alone. The model showed moderate discrimination, and comparison with Framingham or SCORE was not feasible due to missing key variables. Future studies should validate these findings in larger, multi-ethnic cohorts; and incorporate mediation and sensitivity analyses to clarify pathways associating cluster membership to outcomes.

## Ethics Approval Statement

Ethics approval was obtained from the local centralized institutional review board (2015/2603), and all participants provided written informed consent. ClinicalTrials.gov identifier: NCT02670031.

## Data Availability

Data available on request from the corresponding author

## Author’s contribution

TTL and CWLC contribution to the conception and design of the study. AARM, JKC JAB, VL and RB contributed to the acquisition, analysis and interpretation of the data. AARM and JKC drafted the manuscript under the supervision of TTL and CWLC. All authors made critical revisions to the manuscript, gave final approval and agreed to be accountable for all aspects of work ensuring integrity and accuracy.

