## Supplementary material for "Clinical, Prognostic and Biological Features of High-Risk Cardiometabolic Phenotype: the REMODEL Study": Online Supplemental Data

**Cardiovascular Magnetic Resonance**

Cardiovascular magnetic resonance was performed in all participants (Siemens Aera 1.5T, Siemens Healthineers, Erlangen, Germany). Balanced steady-state free precession cine images were acquired in the long-axis (2-, 3- and 4-chamber) and contiguous short-axis cines from the mitral valve annulus to the apex (acquired voxel size 1.6 x 1.3 x 8.0mm; 30 phases per cardiac cycle).

Late gadolinium enhanced imaging was performed 8 minutes following 0.1 mmol/kg of gadobutrol (Gadovist; Bayer Pharma AG, Germany), using an inversion-recovery fast gradient echo sequence. Inversion time was optimized to achieve appropriate nulling of the myocardium. Extracellular volume (ECV) fraction was derived from native and post-contrast myocardial T1 maps, obtained using the modified Look-Locker inversion-recovery sequence. Interstitial volume was calculated as the product of ECV fraction and myocardial volume, while myocyte volume was computed as (1-ECV fraction) x myocardial volume. Myocardial volume (mL) was derived by dividing myocardial mass (g) by the specific density of the myocardium (1.05 g/mL). Values were indexed to body surface area.

**Selection of Features (Predictor Variables)**

Variables with more than 5% missing data were excluded from the analysis. For the remaining variables, missing values were imputed using a k-nearest neighbours (KNN) imputation method. The final dataset consisted of 83 predictors, encompassing demographic (n=9), clinical (n=23), blood-based (n=6), and CMR (n=45) variables. To evaluate the discriminative ability of each predictor for the primary outcome, variables were ranked in descending order based on their individual area under the receiver operating characteristic curve (AUC). Feature selection proceeded sequentially: starting with the top-ranked predictor, additional variables were added in order of decreasing AUC using logistic regression until no further gain in discriminative performance was observed. The final feature set was then reviewed and refined based on clinical relevance to ensure interpretability.

**Determining the Number of Clusters**

The silhouette method was used to evaluate clustering quality by measuring how well an object was assigned to its cluster compared to others^18^ . The optimal number of clusters was determined by selecting the value of k that yielded the highest average silhouette score. A silhouette score close to 1 indicates strong clustering, where an observation is highly similar to others in the same cluster and well differentiated from those in other clusters. A score near 0 suggests that the observation lies near the boundary between clusters, indicating uncertainty in assignment.

**Cluster Assignment using KAMILA**

KAMILA clustering was used to stratify the study cohort based on the selected features and the optimal number of clusters determined above. During iterative optimization, each observation was assigned to the cluster that maximized its likelihood. As cluster membership evolved, KDE estimates were continuously updated to better reflect the distribution of data within each cluster. The algorithm was run with 5,000 random initializations and up to 5,000 iterations per run to ensure robustness and avoid convergence to local optima. This exhaustive strategy promoted stability and reproducibility in the final clustering solution.

**Sensitivity and Stability Analyses for Cluster Selection**

We evaluated candidate solutions for k = 2–10 and computed three complementary validity indices: average silhouette width (using Gower distance), Calinski–Harabasz (CH), and Davies–Bouldin (DB). The k = 2 solution showed the strongest support across indices (silhouette = 0.731, highest; DB = 1.197, lowest), whereas CH peaked at k = 9; using a combined rank approach (silhouette and CH ranked high-to-low; DB low-to-high), k = 2 had the best overall rank-sum and was selected.

We assessed stability using bootstrap subsampling (200 iterations; 80% of observations per iteration) with reclustering via KAMILA and quantified agreement with the baseline using the Adjusted Rand Index (ARI): mean ARI = 0.956 (SD 0.046), with 98% of runs achieving ARI ≥ 0.90; consensus clustering metrics also indicated low ambiguity (PAC (0.1–0.9) = 0.036; WCC = 0.990; BCC = 0.031).

Finally, robustness checks showed partial sensitivity to scaling (ARI vs baseline: z-score 1.00; min–max 0.683; no scaling 0.267; robust scaling 0.216), and broadly consistent recovery of the two-cluster structure using alternative Gower-based methods (PAM and hierarchical clustering; ARI ≈ 0.80), supporting the primary k = 2 finding.

**Proteomics Analysis**

Proteomic profiling was performed using the Olink® Target Cardiovascular Disease II and III panels (Olink Proteomics, Uppsala, Sweden). Protein abundance was reported as normalized protein expression (NPX) values on a log2 scale. Quality control followed Olink recommendations. Samples or assays failing internal controls were excluded, and proteins with low call rates were removed. Values below the limit of detection were retained and replaced with the limit of detection divided by √2.

NPX values were pre-normalized by Olink using internal and inter-plate controls. Residual plate or batch effects were assessed and corrected using empirical Bayes adjustment (ComBat), without inclusion of outcome variables. Proteins with >5% missing values were excluded; remaining missing values were imputed using k-nearest neighbours.

Between-cluster comparisons were performed using two-sample statistical testing, with multiple testing controlled using the Benjamini–Hochberg false discovery rate (FDR). Pathway enrichment analyses were not performed and proteomic findings are hypothesis-generating.
